# Effects of emotional arousal on amygdala neural response and cigarette craving reduction during repeated exposure to graphic warning labels

**DOI:** 10.1101/2025.03.27.25324796

**Authors:** Astrid P. Ramos-Rolón, Daniel D. Langleben, Kevin G. Lynch, Corinde E. Wiers, Zhenhao Shi

## Abstract

Graphic warning labels (GWLs) have been implemented on cigarette packaging worldwide. In the U.S., GWLs have encountered legal obstacles based on the tobacco industry arguments that their aversive imagery unnecessarily triggers strong emotional arousal. This longitudinal study evaluated the effect of the high-arousal GWLs on cigarette craving and the neural substrates of emotional processing. 158 adult smokers were exposed to either high-arousal (n=79) or low-arousal (n=79) GWLs that were attached to their own cigarette packs for 4 weeks. Craving and brain activity in response to GWLs and control stimuli were measured using functional magnetic resonance imaging before and after the 4-week exposure. The amygdala, which plays a key role in emotional processing, served as the a priori region of interest. Results indicate that, at baseline, high-arousal GWLs elicited a larger reduction in craving and stronger amygdala neural response compared to low-arousal GWLs; however, at week 4, GWL-induced craving reduction and amygdala response became comparable between the high-arousal and low-arousal groups. Amygdala response mediated GWLs’ effects on craving reduction, which was moderated by arousal and time in such a way that the amygdala’s mediating role was more pronounced for high-arousal than low-arousal GWLs at baseline but did not differ between groups at week 4. Together, the results suggest that the impact of emotional arousal on cigarette cravings decreases over time, potentially due to the amygdala’s diminishing responsivity to repeated presentations of high-arousal imagery. Low-arousal GWLs may represent a more feasible approach for tobacco control efforts in the U.S.

## INTRODUCTION

Tobacco use is a major public health issue, with over 1.2 billion users worldwide in 2022 (World Health Organization, 2024). Combustible cigarette smoking remains the most prevalent form of tobacco consumption (World Health Organization, 2024). In the past two decades, many countries have implemented graphic warning labels (GWLs) on cigarette packages (Hiilamo et al., 2014). The U.S. first adopted text-only warnings in 1966, and in 2009, Congress tasked the Food and Drug Administration (FDA) with GWL implementation (Hiilamo et al., 2014). Using data from an online survey (Nonnemaker et al., 2010), the FDA selected the GWLs that evoked the highest emotional response (ER) and mandated them in a Final Rule (U.S. Food and Drug Administration, 2011). However, this Final Rule was overturned in court after tobacco companies successfully argued that the GWLs were overly emotionally provocative and infringed on their First Amendment rights to commercial free speech (“R.J. Reynolds Tobacco Company, et al. v. U.S. Food & Drug Administration, et al.,” 2012). In 2020, the FDA introduced a new set of 11 GWLs, which have not been tested for ER (U.S. Food and Drug Administration, 2022). The new GWLs faced similar legal challenges (“R.J. Reynolds Tobacco Company, et al. v. U.S. Food & Drug Administration, et al.,” 2022; “R.J. Reynolds Tobacco Company, et al. v. U.S. Food & Drug Administration, et al.,” 2024b) and were blocked again in January 2025 (“R.J. Reynolds Tobacco Company, et al. v. U.S. Food & Drug Administration, et al.,” 2025).

This debate has spurred research into the risk vs. benefit of using highly emotional imagery in GWLs. Studies show that GWLs evoking a higher ER lead to a greater immediate reduction in cigarette craving and better GWL recall compared to lower-ER GWLs (Wang, Lowen, et al., 2015; Wang, Romer, et al., 2015). However, research to date has largely focused on the short-term effects of GWLs, although in real life, smokers would be exposed to GWLs in a repeated and chronic manner. To bridge this gap, our group examined the longer-term effects of exposure to high-ER vs. low-ER GWLs (Shi, Wang, Fairchild, Aronowitz, Padley, et al., 2023). We found that compared to high-ER GWLs, a month-long exposure to low-ER GWLs was associated with a greater reduction in cigarette consumption. These findings appear to contradict the cross-sectional evidence that postulated greater efficacy of high-ER GWLs largely based on intermediate outcome variables such as recall, attitudes and intentions. Reconciling the cross-sectional and longitudinal findings is crucial for a full mechanistic understanding of the dynamic impact of ER on GWL efficacy.

Craving is a central characteristic of substance use disorders (American Psychiatric Association, 2013) and a key motivational driver of drug seeking (Robinson & Berridge, 1993). Craving for cigarettes robustly precipitates subsequent smoking (Carter et al., 2008; Motschman et al., 2018; Shiffman et al., 2002) and predicts cessation failure (Fidler et al., 2011). While smoking temporarily alleviates craving (Carter et al., 2008), chronic smoking exacerbates craving in the long term which can persist after extended abstinence (Robinson & Berridge, 1993). Reducing craving is an important goal in the clinical and public health interventions of tobacco use (Do & Galván, 2014; Shiffman et al., 2004; Wang, Lowen, et al., 2015; Wang, Romer, et al., 2015). We previously showed that, cross-sectionally, high-ER GWLs produced greater reductions in craving and cue-reactivity than low-ER GWLs (Wang, Lowen, et al., 2015; Wang, Romer, et al., 2015). However, the salience of affective stimuli (e.g., GWLs) decreases after repeated exposure due to habituation of the brain’s emotion circuits (Fischer et al., 2000; Fridriksson et al., 2018; Shi, Wang, Fairchild, Aronowitz, Padley, et al., 2023).

Specifically, using functional magnetic resonance imaging (fMRI), we showed that GWL-induced neural response in the amygdala, a key region for emotional processing (LeDoux, 2007), was initially greater for high-ER GWLs than low-ER GWLs, but the difference was markedly reduced after one month of chronic GWL exposure (Shi, Wang, Fairchild, Aronowitz, Padley, et al., 2023). Such habituation may attenuate high-ER GWLs’ greater efficacy in reducing craving compared to low-ER ones.

The aim of the present study was twofold: to supplement cross-sectional data (Wang, Lowen, et al., 2015) with longitudinal evidence on the relative efficacy of high-ER vs. low-ER GWLs; and to extend our prior research on cigarette consumption (Shi, Wang, Fairchild, Aronowitz, Padley, et al., 2023) by examining the motivational underpinning (i.e., craving) and neural correlates (Wang, Lowen, et al., 2015). Adult smokers who were randomly assigned to the high-ER and low-ER groups underwent fMRI while viewing GWLs and control stimuli and reported momentary craving. The fMRI assessment was repeated after four weeks of naturalistic GWL exposure, as described elsewhere (Shi, Wang, Fairchild, Aronowitz, Padley, et al., 2023). Briefly, exposure was modeled by providing participants with cigarette packs carrying the high-ER or low-ER GWLs.

We hypothesized that, while high-ER GWLs would initially induce greater amygdala response and greater reduction in cigarette craving compared to low-ER GWLs, the impact of ER would diminish after repeated GWL exposure due to emotional habituation (Fischer et al., 2000; Fridriksson et al., 2018; Shi, Wang, Fairchild, Aronowitz, Padley, et al., 2023). We further hypothesized that amygdala neural activity would be associated with craving reduction and mediate the diminishing effect of ER on GWL efficacy.

## MATERIALS AND METHODS

### Participants

168 adult tobacco smokers were recruited from the Philadelphia area and randomly assigned to either the high-ER group or the low-ER group (Shi, Wang, Fairchild, Aronowitz, Padley, et al., 2023). Eight participants were excluded from the current analysis due to missing fMRI data, and two were excluded due to excessive head motion during fMRI, resulting in a total of 158 participants included in the final analysis (n=79 for each group). Inclusion criteria were: 1) 18 to 65 years old; 2) currently smoking ≥2 cigarettes per day; 3) ≥100 cigarettes smoked during lifetime; 4) not currently enrolled in any smoking cessation programs; 5) not currently using other nicotine products (chewing tobacco, cigars, electronic cigarettes, etc.); 6) not a member of the household or friend of an already enrolled study participant; 7) no medical or psychiatric disorders or treatments that may affect the cerebrovascular system; 8) no contraindications for fMRI; 9) (if female) not pregnant or breast-feeding. Participants were instructed to avoid using any other nicotine products throughout the study. All participants provided written informed consent to participate in the protocol approved by the University of Pennsylvania Institutional Review Board.

### Study design

Participants were enrolled seven days before GWL exposure (day –7) and received a week’s supply of cigarettes of their preferred brands in their original packaging without GWLs. Upon enrollment, demographic information and age of smoking initiation were collected, and nicotine dependence was measured by the Fagerström Test for Nicotine Dependence (FTND) (Heatherton et al., 1991). On days 0, 7, 14, and 21, participants received weekly cigarette supply in packages that carried either high-or low-ER GWLs based on their assigned experimental group (see **Figure 1**). To discourage participants from stockpiling the cigarettes, all packages had been unsealed prior to being dispensed. Brain activity and cigarette craving in response to GWLs were measured using fMRI immediately before (day 0) and after (day 28) the four weeks’ GWL exposure. Other measures included the number of cigarettes smoked per day during the past seven days (CPD), urine concentration of nicotine metabolite cotinine, desire to quit smoking, self-efficacy, and perceived GWL effectiveness. Details about these measures and their results were reported in Shi, Wang, Fairchild, Aronowitz, Padley, et al. (2023).

**Figure 1.**
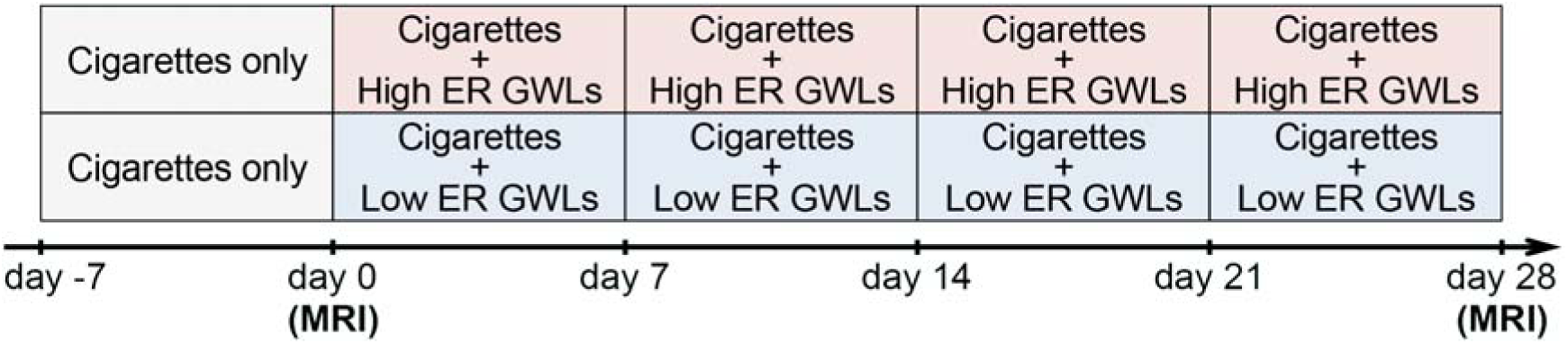
Study timeline schematic.

### Study stimuli

The FDA evaluated the ER to 36 GWLs composed of colored images paired with one of nine textual warnings using an online survey with over 18,000 respondents (Nonnemaker et al., 2010). Out of these 36 GWLs, we selected twelve GWLs with higher ER scores and twelve with lower ER scores (mean±SD=18.21±1.25 versus 16.06±1.09, Mann-Whitney U=207, p=0.001) that carried the same textual warnings and were comparable in terms of image brightness, contrast, color saturation, depiction of human faces, depiction of smoking cues, and usage of photographs as opposed to infographics (ps>0.12) (Shi, Wang, Fairchild, Aronowitz, Padley, et al., 2023). The GWLs covered 50% of the front and back of the packages and were rotated using a Latin square design so that each participant received approximately the same proportion of each specific GWL during the four weeks of exposure.

### fMRI data acquisition

fMRI was performed on a Siemens (Siemens, Erlangen, Germany) TIM Trio 3T system in 50 participants (26 high ER, 24 low ER) and a Siemens Prisma 3T system in 108 participants (53 high ER, 55 low ER; χ²(1)=0.03, p=0.86). We previously showed that inter-scanner variability and its impact on neural activity measures were minimal, allowing us to combine data in subsequent analysis (Shi, Wang, Fairchild, Aronowitz, Padley, et al., 2023). The scanners used identical parameters outlined below. Blood oxygenation level-dependent (BOLD) fMRI used a whole-brain, single-shot gradient-echo echo-planar sequence with the following parameters: repetition time (TR)/echo time (TE)=2000/30 ms, field of view (FOV)=220 mm², matrix=64×64, slice thickness/gap=3.4/0 mm, 32 slices, effective voxel resolution of 3.4×3.4×3.4 mm³. After BOLD fMRI, 5-minute T1-weighted 3D magnetization-prepared gradient-echo structural images were acquired with the following parameters: TR/TE=1810/3.51 ms, FOV=240×180 mm², matrix=256×192, effective voxel resolution of 0.94×0.94×0.94 mm³. An oblique acquisition oriented along the anterior commissure–posterior commissure line allowed coverage of the entire brain except the lower cerebellum.

Participants completed the fMRI task at day 0 and day 28. To ensure that participants were in a non-deprived state during the scans, they were escorted outside approximately 30 minutes before each fMRI session to smoke one of their own cigarettes. The fMRI task followed a block design, with stimuli presented in six blocks of GWLs and six blocks of control images. Each GWL block comprised six of the twelve high-ER or low-ER GWLs, depending on the participant’s group assignment. Each GWL was presented three times throughout the task. Similarly, each control block contained six of twelve smoking-irrelevant control images (e.g., buildings) paired with smoking-irrelevant text (e.g., “WARNING: Buildings are hazardous”), matching the anti-smoking text warnings on the GWLs in terms of word count, font style, and font size. Each GWL and image was presented for 2 seconds against a black background, resulting in a block duration of 12 seconds. Immediately before and after each block, participants were asked to indicate their current level of cigarette craving by answering the question “How much do you want to smoke a cigarette right now?” using a visual analog scale from 0 (“not at all”) to 10 (“extremely”). They were given 4 seconds to provide their response. Inter-block intervals, indicated by a white crosshair in the center of the screen, lasted for 9.5 seconds. All stimuli were delivered using Presentation (Neurobehavioral System Inc, Albany, CA) and presented using a rear projector system that was viewed through a mirror mounted on the head coil in the scanner.

### fMRI data processing

fMRI data were analyzed using SPM 12 (Wellcome Trust Centre for Neuroimaging, London, UK). Images were adjusted for slice timing, realigned to the first scan to correct for head motion, normalized into stereotactic Montreal Neurological Institute (MNI) space with 3-mm isotropic cubic voxels, and spatially smoothed by a Gaussian filter with full-width at half-maximum parameter set to 8 mm. Severe head motion was defined as motion greater than one voxel, a commonly used cutoff in the task-fMRI literature. Such a cutoff ensured the robustness of SPM’s default motion correction algorithm (spm_realign.m) (Friston et al., 1995; Friston et al., 1996). The root-mean-squared framewise displacement of the remaining participants was minimal (range=0.02– 0.35 mm) (Jenkinson et al., 2002).

After preprocessing, we conducted individual-level general linear model (GLM) analysis using the “beta series” approach (Zanto et al., 2011) to model each block with the canonical hemodynamic response function as a single regressor. High-pass filter of 128 sec was applied. Six rigid body motion parameters (x, y, z, pitch, roll, yaw) and their first-order derivatives were included as covariates, and the rest periods were treated as the implicit baseline. The GLMs resulted in a total of twelve parameter-estimate maps for each participant and each fMRI session (six for the GWL blocks and six for the control image blocks). The bilateral amygdala was anatomically defined as an a priori regions of interest using the Automated Anatomical Labeling atlas (Tzourio-Mazoyer et al., 2002). For each participant and each fMRI session, the percent signal change in the amygdala in response to each GWL and control block was extracted for subsequent statistical analysis.

### Statistical analysis

Statistical analysis was performed using R. We compared baseline participant characteristics between the high-ER and low-ER groups using χ² tests for categorical variables and Mann-Whitney U-tests for numerical variables. We then evaluated the external validity of the craving measure. To do so, we examined the association of mean craving with nicotine dependence measured by FTND and with smoking severity indexed by CPD. Mean craving was calculated by averaging all 24 craving ratings (i.e., before and after the six GWL and six control image blocks) for each subject and each fMRI session. CPD was log-10 transformed due to high right-skewness. A linear model examined the effect of nicotine dependence on mean craving at day 0 while controlling for group (i.e., mean_craving ∼ 1 + group + FTND). A linear mixed-effects (LME) model examined the effect of CPD on mean craving across both the day-0 and day-28 time points with random subject-specific intercepts while controlling for time and group [i.e., mean_craving ∼ 1 + time + group + CPD + (1|subject)]. LME analysis was performed using the lme4 and lmerTest packages (Bates et al., 2015; Luke, 2017). Statistical significance was evaluated with the Satterthwaite approximation for degrees of freedom (Luke, 2017).

The effect of GWLs and control images on cigarette craving during the fMRI task was calculated as the change in craving ratings from before to after each GWL block and each control image block, respectively. Therefore, there was a total of twelve scores of craving reduction (i.e., Δcraving) for each participant and each fMRI session (six for the GWL blocks and six for the control image blocks). Similarly, twelve values of amygdala percent signal change were extracted per participant per session. A series of LME models were conducted to test for the study hypotheses. For all models, random intercepts were included to account for repeated measures across GWL and control image blocks within subjects and subject-time combinations. We first tested the diminishing role of ER in GWLs’ ability to reduce cigarette craving after repeated exposure. Specifically, the model examined the effects of time (day 0 vs. day 28), group (high-ER vs. low-ER), and stimulus (GWL vs. control), on smoking reduction [i.e., the *c*_(•)_ paths in **Figure 5B**: Δcraving ∼ 1 + time*group*stimulus + (1|subject) + (1|subject:time)]. Post-hoc analysis was conducted to decompose three-way interactions using the emmeans package, and multiple comparisons were adjusted using the Šidák method. For completeness and as a replication of our previously reported analysis (Shi, Wang, Fairchild, Aronowitz, Padley, et al., 2023), a similar LME model was used to examine the effects of time, group, and stimulus on amygdala neural activity [i.e., the *a*_(•)_ paths in **Figure 5B**: amygdala ∼ 1 + time*group*stimulus + (1|subject) + (1|subject:time)]. Next, we tested the association between amygdala response and craving reduction [i.e., the *b* paths in **Figure 5B**: Δcraving ∼ 1 + amygdala + (1|subject) + (1|subject:time)].

Lastly, we tested whether amygdala response mediated high-ER GWLs’ decreasing ability to reduce craving after repeated exposure. We first tested an unmoderated mediation model, where stimulus was the input variable, amygdala response was the mediator, and craving reduction was the outcome. We then tested a moderated mediation model adapted from Hayes (2015) with time and group being the moderators (see **Figure 5**). Both models involved an LME analysis for the mediator (i.e., the *a*_(•)_ paths) and another LME analysis for the outcome (i.e., the *b* and *c*_(•)_ paths combined). The index of moderated mediation was calculated as *a*_7_×*b*. The conditional indirect effect of stimulus on craving through amygdala activity, which varied as a function of time and group, was calculated as (*a*_3_ + *a*_5_×time + *a*_6_×group + *a*_7_×time×group)×*b*. Bootstrap using case resampling (10000 iterations) (Reiser et al., 2017) was performed to determine the bias-corrected and accelerated confidence interval and significance of effects.

## RESULTS

### Participant characteristics

Participants of the high-ER and low-ER groups were comparable with regard to demographics and baseline characteristics (see **Table 1**). All participants completed the first fMRI session, and 140 participants (89%) completed the second fMRI session. The completion rate for the second fMRI session was comparable between the high-ER and low-ER groups (n=71 vs. 69, χ²(1)=0.06, p=0.80).

**Table 1.** Participant characteristics at baseline (count or mean±SD).

| Variable | High-ER (n=79) | Low-ER (n=79) | p-value<sup>a</sup> |
| --- | --- | --- | --- |
| Age | 29.4±9.6 | 27.8±8.0 | 0.42 |
| Sex | 37 female, 42 male | 38 female, 41 male | >0.99 |
| Race | 44 White, 20 AA,<br>6 Asian, 9 other | 44 White, 18 AA,<br>6 Asian, 11 other | 0.96 |
| Ethnicity | 11 Hispanic | 5 Hispanic | 0.19 |
| Age of smoking initiation (years) | 16.5±3.2 | 17.6±4.4 | 0.099 |
| Nicotine dependence (FTND)<sup>b</sup> | 4.6±2.6 | 4.3±2.4 | 0.39 |
| Number of cigarettes smoked per day<sup>b</sup> | 13.5±8.6 | 12.0±6.6 | 0.44 |

Abbreviations: ER, emotional response; FTND, Fagerström Test for Nicotine Dependence; AA, African American.
<sup>a</sup> P-values were obtained from group comparisons using $\chi^2$ tests for categorical variables and Mann-Whitney U-tests for numerical variables.
<sup>b</sup> Baseline FTND and number of cigarettes was missing in two and one participant, respectively.

### External validity of craving

Mean cigarette craving across all ratings during the MRI session at day 0 was positively associated with baseline nicotine dependence indexed by FTND (β=0.22, 95% confidence interval [CI]=[0.06,0.39]; F(1,153)=7.18, p=0.008). Mean cigarette craving was also positively associated with smoking severity indexed by CPD across day 0 and day 28 (β=2.21, 95% CI=[0.88,3.53]; F(1,205.34)=10.61, p=0.001). There was a trend for a main effect of time on mean craving, with marginally higher mean craving at day 0 than day 28 (4.12 vs. 3.71, difference=0.41, 95% CI=[–0.003,0.83]). The main effect of group and the time×group interaction were not significant (p=0.68 & 0.78).

### Effect of GWLs on craving

There was a significant time×group×stimulus interaction on craving reduction (F(1,2919.96)=9.944, p=0.002) (see **Figure 2**). Post-hoc analysis showed significant group×stimulus interaction at day 0 (p<0.001), such that the reduction of craving induced by the GWLs (vs. control images) was greater in the high-ER group (–0.81 vs. 0.37, difference=–1.18, 95% CI=[–1.44,–0.92]) than the low-ER group (–0.50 vs. 0.09, difference =–0.59, 95% CI=[–0.85,–0.33]). The group×stimulus interaction was not significant at day 28 (p=0.86), with comparable effect of GWLs vs. control images on the reduction of craving between the high-ER group (–0.38 vs. 0.00, difference=–0.38, 95% CI=[–0.65,–0.12]) and the low-ER group (–0.49 vs. –0.04, difference=–0.45, 95% CI=[–0.72,–0.19]).

**Figure 2.**
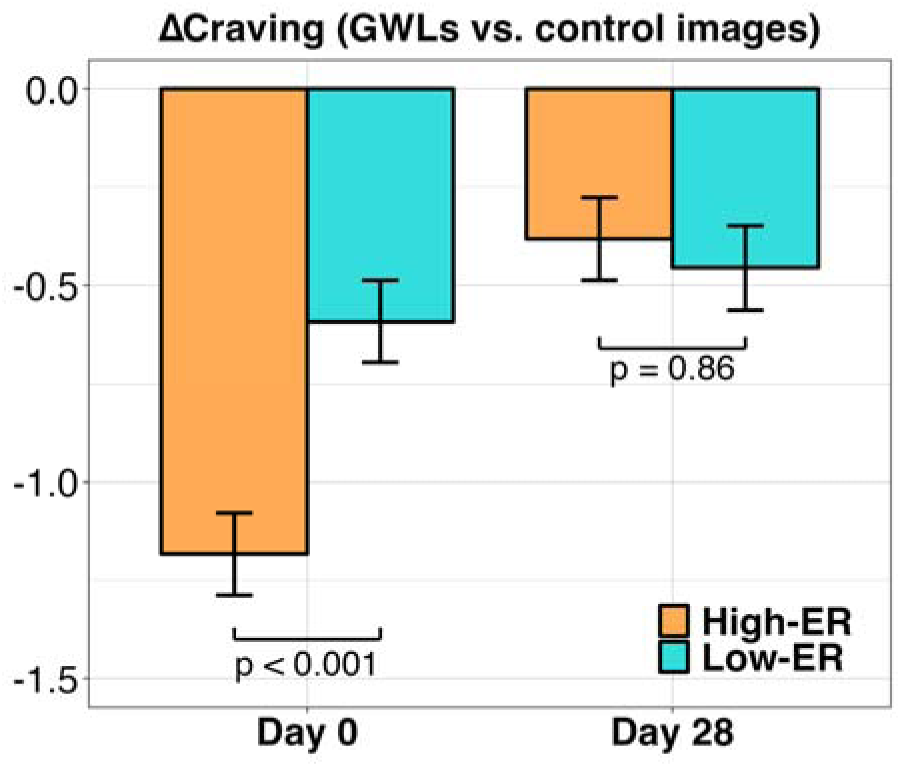
Difference in cigarette craving reduction between GWL blocks and control image blocks across time (day 0 vs. day 28) and group (high-ER vs. low-ER). Abbreviations: GWL, graphic warning label; ER, emotional response.

### Effect of GWLs on amygdala neural activity

There was a significant time×group×stimulus interaction on amygdala neural activity (F(1,3274)=5.36, p=0.021) (see **Figure 3**). Post-hoc analysis showed significant group×stimulus interaction at day 0 (p<0.001), such that amygdala response to GWLs (vs. control images) was greater in the high-ER group (0.18 vs. –0.03, difference=0.21, 95% CI=[0.13,0.29]) than the low-ER group (0.04 vs. 0.02, difference=0.03, 95% CI=[– 0.05,0.10]). The group×stimulus interaction was not significant at day 28 (p=0.69), with comparable amygdala response to GWLs vs. control images between the high-ER group (0.26 vs. 0.13, difference=0.13, 95% CI=[0.04,0.21]) and the low-ER group (0.08 vs. –0.01, difference=0.09, 95% CI=[0.01,0.17]). These results were consistent with the original analysis reported in Shi, Wang, Fairchild, Aronowitz, Padley, et al. (2023).

**Figure 3.**
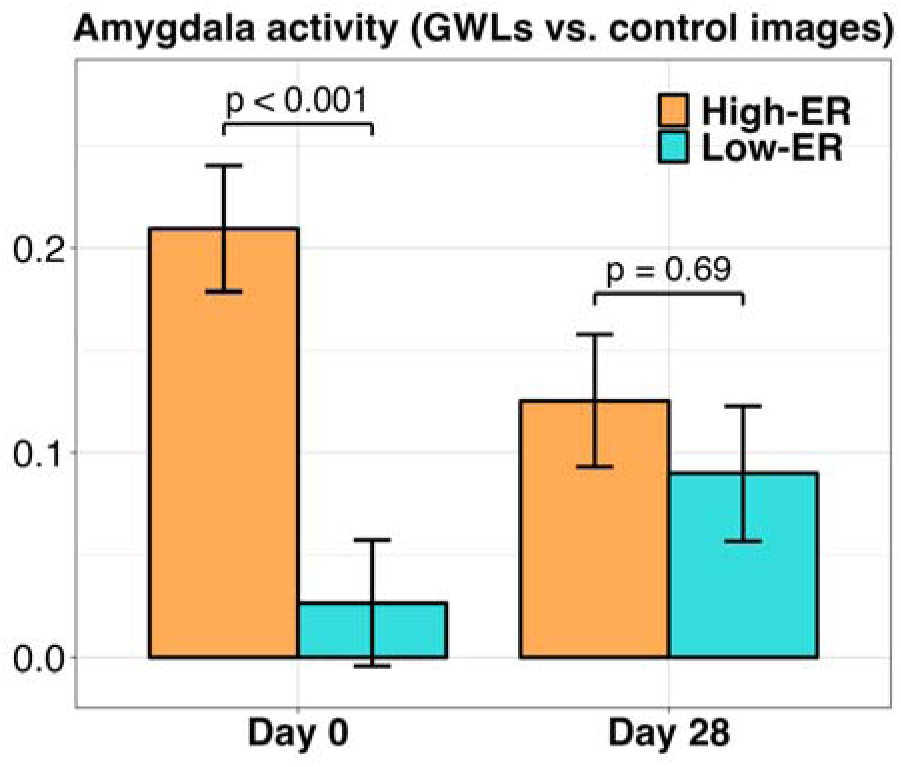
Amygdala activity in response to GWLs vs. control images across time (day 0 vs. day 28) and group (high-ER vs. low-ER). Abbreviations: GWL, graphic warning label; ER, emotional response.

### Association between craving reduction and amygdala neural activity

Greater amygdala response was associated with greater reduction in cigarette craving (β=–0.15, 95% CI=[– 0.22,–0.08]; F(1,703.36)=16.79, p<0.001) (see **Figure 4**). The association remained significant when controlling for the effects of time, group, stimulus, and their interactions (β=–0.11,95% CI=[–0.19,–0.04]; F(1,721.39)=9.56, p=0.002).

**Figure 4.**
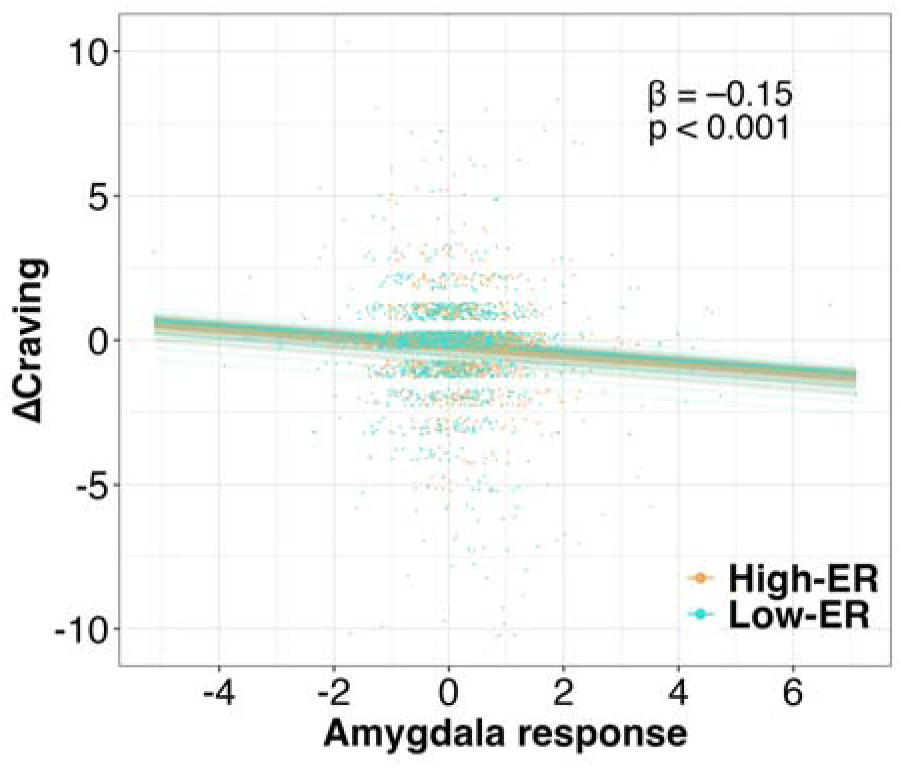
Significant association between change in cigarette craving and amygdala neural activity. Abbreviation: ER, emotional response.

### Mediation analysis

We found a significant unmoderated indirect effect of stimulus on craving reduction through amygdala neural activity (effect=−0.006, 95% CI=[–0.01,–0.001]; p=0.022), suggesting a key role of amygdala in mediating GWLs’ impact on craving. We then examined the moderated mediation model, as shown in **Figure 5**. We found a significant moderated mediation, such that the mediator effect was moderated by the group×time interaction (effect=−0.002, 95% CI=[–0.005,–0.0001]; p=0.035) (see **Figure 6**). Post-hoc analysis showed that, at day 0, the indirect effect of stimulus on craving reduction through amygdala activity was stronger for the high-ER group (β=−0.011, 95% CI=[–0.021,–0.001]) than the low-ER group (β=−0.0004, 95% CI=[–0.005,0.003]; p=0.020). At day 28, the indirect effect was comparable between the high-ER group (β=−0.007, 95% CI=[–0.018,–0.001]) and the low-ER group (β=−0.005, 95% CI=[–0.013,–0.0005]; p=0.41).

**Figure 5.**
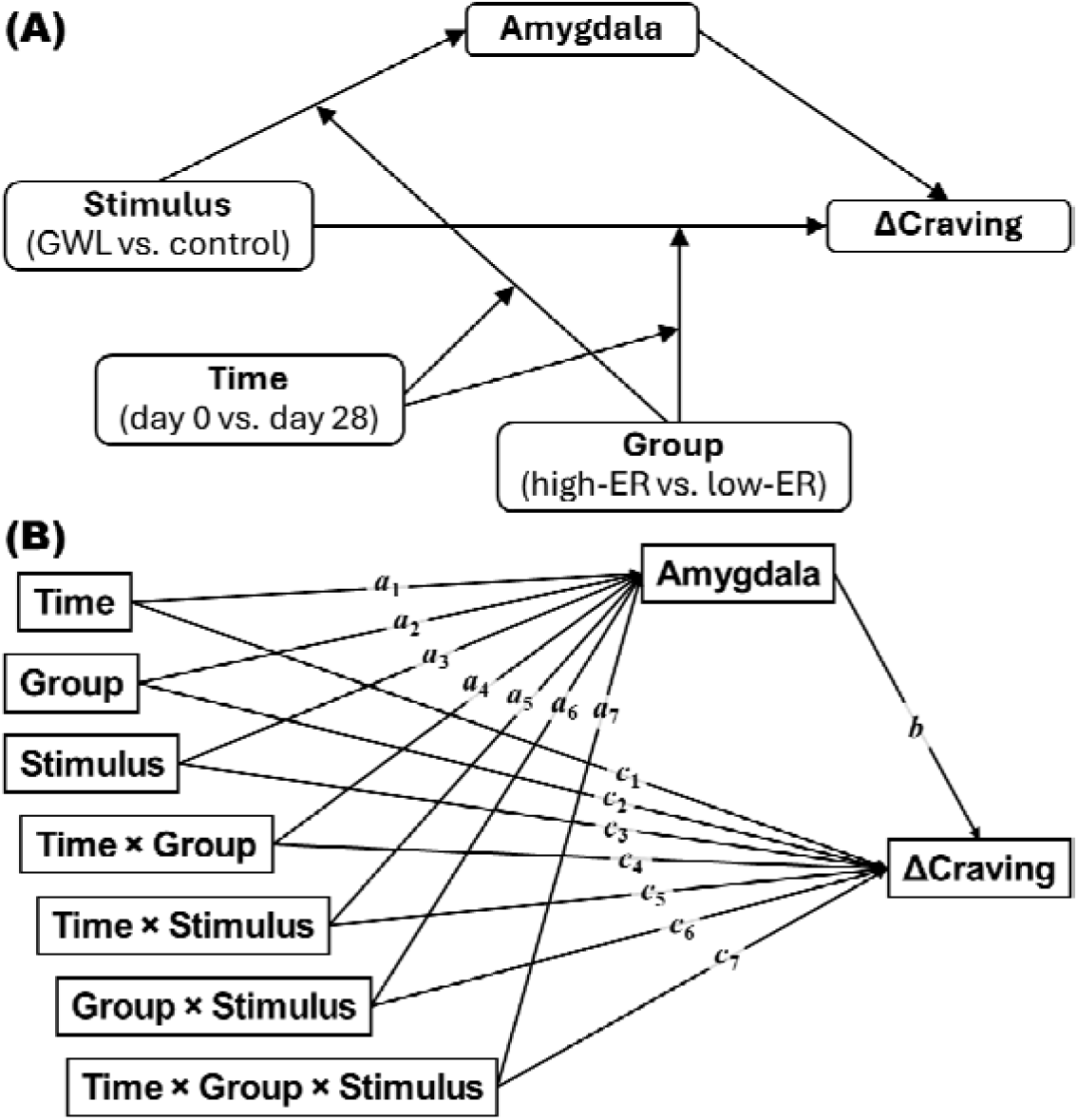
Moderated mediation model presented in the conceptual form (A) and the statistical form (B) (adapted from Hayes (2015)).

**Figure 6.**
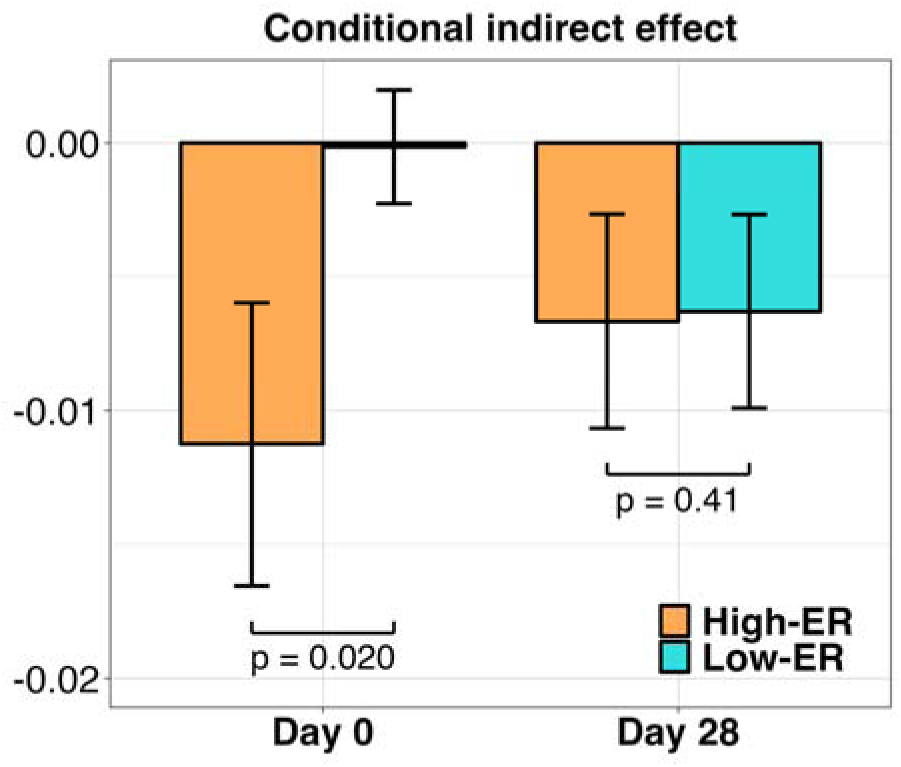
Conditional indirect effect of stimulus (GWLs vs. control images) on Δcraving through amygdala neural activity across time (day 0 vs. day 28) and group (high-ER vs. low-ER). Abbreviations: GWL, graphic warning label; ER, emotional response.

## DISCUSSION

We found that high-ER GWLs initially elicited a stronger amygdala response and greater craving reduction compared to low-ER GWLs, similar to what we observed in our prior cross-sectional study (Wang, Lowen, et al., 2015; Wang, Romer, et al., 2015). However, these differences diminished with repeated GWL exposure, such that by day 28, amygdala response and craving reduction became comparable between the high-ER and low-ER groups. Mediation analyses further revealed that amygdala response mediated the effects of GWLs on craving reduction. The mediating effect was stronger for high-ER than low-ER GWLs at baseline but not at day 28.

The diminishing effect of ER on amygdala response is consistent with emotional habituation, a process in which an individual’s reaction to repeatedly presented affective stimuli gradually decreases over time. While our data demonstrated amygdala habituation over four weeks of chronic GWL exposure, previous studies have reported more rapid amygdala adaptation occurring within seconds when identical GWLs were presented consecutively (Fridriksson et al., 2018). Habituation is a fundamental form of learning that optimizes cognitive resource allocation and facilitates the differentiation between familiar and novel stimuli. fMRI studies have specifically shown amygdala habituation to emotional stimuli across various modalities, including visual (Breiter et al., 1996; Plichta et al., 2014; Wright et al., 2001), auditory (Mutschler et al., 2010), and olfactory (Poellinger et al., 2001). Despite the decreasing amygdala activity, we previously showed that high-ER GWLs were better remembered over time (Wang et al., 2019). This indicates that while amygdala-mediated emotional processing may facilitate encoding, it may not be necessary for retention. Taken together, our findings highlight the importance of considering emotional habituation in the design of anti-smoking warnings, such as GWLs, that are intended for prolonged exposure among smokers. Strategies to mitigate habituation, such as reducing exposure frequency and increasing stimulus diversity, have shown promise; however, their effectiveness also wanes over time (Rankin et al., 2009). Further research is needed to evaluate the utility and feasibility of alternative approaches for enhancing GWLs’ ability to emotionally engage smokers in the long term.

In addition to habituation, prolonged exposure may trigger reactance, a psychological resistance to perceived persuasion attempts, leading smokers to consider the warnings as threatening and thus dismiss or counterargue them (Brehm & Brehm, 2013). GWLs have been shown to lead to reactance more so than textual anti-smoking warnings (Erceg-Hurn & Steed, 2011; LaVoie et al., 2015; Maynard et al., 2014). Reactance to GWLs has been associated with reduced intention to quit smoking and more cigarettes smoked per day (Hall et al., 2018; Hall et al., 2017) [but see Cho et al. (2016)]. Moreover, chronic GWL exposure may also lead to message fatigue, where individuals become tired of seeing the warnings and perceive them as annoying and uninformative (So et al., 2017). Over time, message fatigue can result in reduced attention, lower emotional engagement, and decreased motivation to change behaviors (Keating & Skurka, in press). Our finding that high-ER GWLs’ ability to reduce craving diminished over time suggest that ER may contribute to reactance and message fatigue elicited by GWLs. Although we did not explicitly measure reactance and message fatigue, our recent exploratory analysis of the same dataset showed that, after completing the four weeks’ GWL exposure, smokers expressed a moderate sense of relief from having to see the GWLs (Shi et al., 2024). While such sense of relief did not differ between the high-ER and low-ER groups, it was only within the high-ER group that greater relief predicted less smoking (Shi et al., 2024). Therefore, addressing reactance and message fatigue may potentially help enhance the behavioral efficacy of high-ER GWLs.

Our previous work examined GWL effects on smoking behavior (Shi, Wang, Fairchild, Aronowitz, Padley, et al., 2023), while the current study focused on a key motivational underpinning of smoking, i.e., craving. Both studies indicate that low-ER GWLs are non-inferior to high-ER GWLs during chronic naturalistic exposure. However, while we previously showed that low-ER GWLs was more effective in reducing smoking (indexed by the number cigarettes per day), here we found comparable efficacy between high-ER and low-ER GWLs in reducing craving after the exposure, despite high-ER ones being more effective initially. Several factors may explain the differences in the two studies’ findings. Smoking and craving function on different time scales— while smoking behavior is a discrete, observable action that unfolds over a longer period (e.g., minutes or hours), craving is a fluctuating internal state that can change moment to moment. The temporal resolution of fMRI (approximately one second) allowed us to integrate momentary assessments of craving in the fMRI task and measure its association with neural activity, providing insight into the immediate psychological and neurobiological effects of GWLs. Beyond craving, smoking behavior is also shaped by a range of additional factors, such as cigarette availability, environmental constraints (Pentz et al., 1989), nicotine metabolism (Pianezza et al., 1998), stress (McKee et al., 2010), and social influence (East et al., 2021). These factors may potentially add to or interact with the effects of craving on actual smoking during GWL exposure (Childs & de Wit, 2010; Dimoff et al., 2019; Gass et al., 2014). Understanding these complex, interrelated factors will be crucial for interpreting the findings of our previous (ref) and current studies and will refine future interventions aimed at reducing smoking through GWLs.

Craving is a multi-dimensional phenomenon involving cognitive, affective, sensory, and physiological components (Kober & DeLeone, 2011; Miele et al., 2023; Rosenberg, 2009). It is one of the eleven diagnostic criteria for tobacco use disorder in DSM-5 (American Psychiatric Association, 2013). Despite some criticisms over the accuracy of self-report measures of craving (Sayette et al., 2000), we found that average craving was positively correlated with nicotine dependence and smoking severity, supporting the external validity of the measure. Factors that contribute to cigarette craving include stress (Childs & de Wit, 2010), nicotine withdrawal (Hughes & Hatsukami, 1986), exposure to smoking-related conditioned stimuli (i.e., cues) (Betts et al., 2021; Shi, Wang, Fairchild, Aronowitz, Lynch, et al., 2023), and spontaneous smoking-related thoughts (e.g., intrusive thoughts and imaginations of cigarette smell) (Aslan et al., 2023). Our previous cross-sectional study showed greater reduction in smoking cue-induced craving after viewing high-ER GWLs compared to low-ER ones (Wang, Romer, et al., 2015). In the current study, craving was measured without the presentation of smoking cues and without the induction of stress or withdrawal. Therefore, our current data reflect diminishing utility of high-ER GWLs in reducing basal craving, which, compared to cue-induced craving, was shown to be more closely related to nicotine dependence and smoking severity (Dunbar et al., 2014). Nevertheless, it would be interesting for future studies to examine whether our longitudinal findings can be generalized to GWL effects on cue-, withdrawal-, and stress-induced craving.

Our study comes with a number or limitations. Firstly, the four-week exposure period, though substantial for neuroimaging, is brief compared to potentially years of exposure to GWLs if implemented. Future studies should extend this timeframe and incorporate real-world measures, such as ecological momentary assessment, to better understand the dynamic and long-term impact of GWLs on craving and smoking outside fMRI. Secondly, given our focus on emotional processing and the amygdala’s well-established role in this domain (LeDoux, 2007), we selected the amygdala as our sole region of interest. Future studies should examine other brain regions and their interregional connectivity, which may further account for GWLs’ efficacy. Thirdly, our study participants were exclusive combustible cigarette smokers who were not currently using other nicotine products. However, the number of electronic cigarette users has been rapidly increasing (Vahratian et al., 2025), and the dual use of electronic and combustible cigarettes has become highly prevalent (Coleman et al., 2022). Examining high-ER and low-ER GWLs’ efficacy in the dual user population would be important for a comprehensive understanding of their public health impact.

The major argument against GWLs used by the tobacco companies in court has been that the highly aversive images included in the GWLs “do not convey any warning information at all … [and] are unabashed attempts to evoke emotion” (“R.J. Reynolds Tobacco Company, et al. v. U.S. Food & Drug Administration, et al.,” 2012), that the GWLs “are designed to shock rather than inform”, and that the “graphics go far beyond what would be necessary to communicate a simple factual message” (“R.J. Reynolds Tobacco Company, et al. v. U.S. Food & Drug Administration, et al.,” 2024a). Our past studies provide both behavioral and neurobiological evidence supporting the effectiveness of GWLs. Notably, we found that under prolonged, real-world exposure conditions, low-ER GWLs are non-inferior to high-ER GWLs in reducing smoking motivation and behavior. Thus, our findings do not support maximizing the ER in GWLs for the greatest public health benefit and suggest that any future GWLs should be pre-tested for ER and their neurobehavioral effects prior to implementation. These findings offer a potential pathway to balancing public health objectives with the constitutional concerns surrounding GWL enforcement.

## DATA AVAILABILITY

The data underlying this article can be shared on reasonable request to the corresponding author.

## AUTHOR CONTRIBUTIONS

**Astrid P. Ramos-Rolón**: formal analysis (equal), investigation (equal), methodology (equal), software (equal), validation (equal), visualization (equal), original draft preparation (equal), review & editing (equal). **Daniel D. Langleben**: conceptualization (lead), data curation (lead), funding acquisition (lead), investigation (equal), methodology (supportive), project administration (lead), resources (lead), supervision (lead), review & editing (equal). **Kevin G. Lynch**: formal analysis (supportive), investigation (supportive), methodology (supportive), software (supportive), review & editing (equal). **Corinde E. Wiers**: conceptualization (supportive), investigation (supportive), methodology (supportive), resources (supportive), original draft preparation (supportive), review & editing (equal). **Zhenhao Shi**: conceptualization (supportive), data curation (supportive), formal analysis (equal), investigation (equal), methodology (equal), project administration (supportive), resources (supportive), software (equal), supervision (supportive), validation (equal), visualization (equal), original draft preparation (equal), review & editing (equal).

## REFERENCES

1. American Psychiatric Association. (2013). Diagnostic and Statistical Manual of Mental Disorders (DSM-5®). American Psychiatric Association.

2. Aslan, M., Sala, M., Gueorguieva, R., & Garrison, K. A. (2023). A network analysis of cigarette craving. Nicotine & Tobacco Research, 25(6), 1155–1163. 10.1093/ntr/ntad021

3. Bates, D., Mächler, M., Bolker, B., & Walker, S. (2015). Fitting linear mixed-effects models using lme4. Journal of Statistical Software, 67(1), 1–48. 10.18637/jss.v067.i01

4. Betts, J. M., Dowd, A. N., Forney, M., Hetelekides, E., & Tiffany, S. T. (2021). A meta-analysis of cue reactivity in tobacco cigarette smokers. Nicotine & Tobacco Research, 23(2), 249–258. 10.1093/ntr/ntaa147

5. Brehm, S. S., & Brehm, J. W. (2013). Psychological Reactance: A Theory of Freedom and Control. Academic Press.

6. Breiter, H. C., Etcoff, N. L., Whalen, P. J., Kennedy, W. A., Rauch, S. L., Buckner, R. L., Strauss, M. M., Hyman, S. E., & Rosen, B. R. (1996). Response and habituation of the human amygdala during visual processing of facial expression. Neuron, 17(5), 875–887. 10.1016/S0896-6273(00)80219-6

7. Carter, B., Lam, C., Robinson, J., Paris, M., Waters, A., Wetter, D., & Cinciripini, P. (2008). Real-time craving and mood assessments before and after smoking. Nicotine & Tobacco Research, 10(7), 1165–1169. 10.1080/14622200802163084

8. Childs, E., & de Wit, H. (2010). Effects of acute psychosocial stress on cigarette craving and smoking. Nicotine & Tobacco Research, 12(4), 449–453. 10.1093/ntr/ntp214

9. Cho, Y. J., Thrasher, J. F., Swayampakala, K., Yong, H.-H., McKeever, R., Hammond, D., Anshari, D., Cummings, K. M., & Borland, R. (2016). Does reactance against cigarette warning labels matter? Warning label responses and downstream smoking cessation amongst adult smokers in Australia, Canada, Mexico and the United States. PLOS One, 11(7), e0159245. 10.1371/journal.pone.0159245

10. Coleman, S. R. M., Piper, M. E., Byron, M. J., & Bold, K. W. (2022). Dual use of combustible cigarettes and e-cigarettes: a narrative review of current evidence. Current Addiction Reports, 9(4), 353–362. 10.1007/s40429-022-00448-1

11. Dimoff, J. D., Sayette, M. A., & Levine, J. M. (2019). Experiencing cigarette craving with a friend: a shared reality analysis. Psychology of Addictive Behaviors, 33(8), 721–729. 10.1037/adb0000519

12. Do, K. T., & Galván, A. (2014). FDA cigarette warning labels lower craving and elicit frontoinsular activation in adolescent smokers. Social Cognitive and Affective Neuroscience, 10(11), 1484–1496. 10.1093/scan/nsv038

13. Dunbar, M. S., Shiffman, S., Kirchner, T. R., Tindle, H. A., & Scholl, S. M. (2014). Nicotine dependence, “background” and cue-induced craving and smoking in the laboratory. Drug and Alcohol Dependence, 142, 197–203.

14. East, K., McNeill, A., Thrasher, J. F., & Hitchman, S. C. (2021). Social norms as a predictor of smoking uptake among youth: a systematic review, meta-analysis and meta-regression of prospective cohort studies. Addiction, 116(11), 2953–2967.

15. Erceg-Hurn, D. M., & Steed, L. G. (2011). Does exposure to cigarette health warnings elicit psychological reactance in smokers? Journal of Applied Social Psychology, 41(1), 219–237.

16. Fidler, J. A., Shahab, L., & West, R. (2011). Strength of urges to smoke as a measure of severity of cigarette dependence: comparison with the Fagerström Test for Nicotine Dependence and its components. Addiction, 106(3), 631–638. 10.1111/j.1360-0443.2010.03226.x

17. Fischer, H. Ê., Furmark, T., Wik, G., & Fredrikson, M. (2000). Brain representation of habituation to repeated complex visual stimulation studied with PET. NeuroReport, 11(1), 123–126.

18. Fridriksson, J. F., Rorden, C., Newman-Norlund, R. D., Froeliger, B., & Thrasher, J. F. (2018). Smokers’ neurological responses to novel and repeated health warning labels (HWLs) from cigarette packages. Frontiers in Psychiatry, 9, 319.

19. Friston, K. J., Ashburner, J., Frith, C. D., Poline, J. B., Heather, J. D., & Frackowiak, R. S. J. (1995). Spatial registration and normalization of images. Human Brain Mapping, 3(3), 165–189.

20. Friston, K. J., Williams, S., Howard, R., Frackowiak, R. S. J., & Turner, R. (1996). Movement-related effects in fMRI time-series. Magnetic Resonance in Medicine, 35(3), 346–355.

21. Gass, J. C., Motschman, C. A., & Tiffany, S. T. (2014). The relationship between craving and tobacco use behavior in laboratory studies: a meta-analysis. Psychology of Addictive Behaviors, 28(4), 1162–1176. 10.1037/a0036879

22. Hall, M. G., Sheeran, P., Noar, S. M., Boynton, M. H., Ribisl, K. M., Parada Jr, H., Johnson, T. O., & Brewer, N. T. (2018). Negative affect, message reactance and perceived risk: how do pictorial cigarette pack warnings change quit intentions? Tobacco Control, 27(e2), e136–e142. 10.1136/tobaccocontrol-2017-053972

23. Hall, M. G., Sheeran, P., Noar, S. M., Ribisl, K. M., Boynton, M. H., & Brewer, N. T. (2017). A brief measure of reactance to health warnings. Journal of Behavioral Medicine, 40(3), 520–529. 10.1007/s10865-016-9821-z

24. Hayes, A. F. (2015). An index and test of linear moderated mediation. Multivariate Behavioral Research, 50(1), 1–22. 10.1080/00273171.2014.962683

25. Heatherton, T. F., Kozlowski, L. T., Frecker, R. C., & Fagerstrom, K. O. (1991). The Fagerstrom Test for Nicotine Dependence: a revision of the Fagerstrom Tolerance Questionnaire. British Journal of Addiction, 86(9), 1119–1127.

26. Hiilamo, H., Crosbie, E., & Glantz, S. A. (2014). The evolution of health warning labels on cigarette packs: the role of precedents, and tobacco industry strategies to block diffusion. Tobacco Control, 23(1), e2. 10.1136/tobaccocontrol-2012-050541

27. Hughes, J. R., & Hatsukami, D. (1986). Signs and symptoms of tobacco withdrawal. Archives of General Psychiatry, 43(3), 289–294. 10.1001/archpsyc.1986.01800030107013

28. Jenkinson, M., Bannister, P., Brady, M., & Smith, S. (2002). Improved optimization for the robust and accurate linear registration and motion correction of brain images. NeuroImage, 17(2), 825–841.

29. Keating, D. M., & Skurka, C. (in press). Meta-analytic evidence that message fatigue is associated with unintended persuasive outcomes. Communication Research. 10.1177/00936502241287875

30. Kober, H., & DeLeone, C. M. (2011). Smoking and neuroimaging: a review. Current Cardiovascular Risk Reports, 5(6), 484–491. 10.1007/s12170-011-0201-5

31. LaVoie, N. R., Quick, B. L., Riles, J. M., & Lambert, N. J. (2015). Are graphic cigarette warning labels an effective message strategy? A test of psychological reactance theory and source appraisal. Communication Research, 44(3), 416–436. 10.1177/0093650215609669

32. LeDoux, J. (2007). The amygdala. Current Biology, 17(20), R868–R874.

33. Luke, S. G. (2017). Evaluating significance in linear mixed-effects models in R. Behavior Research Methods, 49(4), 1494–1502. 10.3758/s13428-016-0809-y

34. Maynard, O. M., Attwood, A., O’Brien, L., Brooks, S., Hedge, C., Leonards, U., & Munafò, M. R. (2014). Avoidance of cigarette pack health warnings among regular cigarette smokers. Drug and Alcohol Dependence, 136, 170–174.

35. McKee, S. A., Sinha, R., Weinberger, A. H., Sofuoglu, M., Harrison, E. L. R., Lavery, M., & Wanzer, J. (2010). Stress decreases the ability to resist smoking and potentiates smoking intensity and reward. Journal of Psychopharmacology, 25(4), 490–502. 10.1177/0269881110376694

36. Miele, C., Cabé, J., Cabé, N., Bertsch, I., Brousse, G., Pereira, B., Moulin, V., & Barrault, S. (2023). Measuring craving: a systematic review and mapping of assessment instruments. What about sexual craving? Addiction, 118(12), 2277–2314. 10.1111/ADD.16287

37. Motschman, C. A., Germeroth, L. J., & Tiffany, S. T. (2018). Momentary changes in craving predict smoking lapse behavior: a laboratory study. Psychopharmacology, 235(7), 2001–2012. 10.1007/s00213-018-4898-4

38. Mutschler, I., Wieckhorst, B., Speck, O., Schulze-Bonhage, A., Hennig, J., Seifritz, E., & Ball, T. (2010). Time scales of auditory habituation in the amygdala and cerebral cortex. Cerebral Cortex, 20(11), 2531–2539. 10.1093/cercor/bhq001

39. Nonnemaker, J., Farrelly, M., Kamyab, K., Busey, A., & Mann, N. (2010). Experimental study of graphic cigarette warning labels (Contract No. HHSF-223-2009-10135G, RTI Project No. 0212305.007.003).

40. Pentz, M. A., Brannon, B. R., Charlin, V. L., Barrett, E. J., MacKinnon, D. P., & Flay, B. R. (1989). The power of policy: the relationship of smoking policy to adolescent smoking. American Journal of Public Health, 79(7), 857–862. 10.2105/AJPH.79.7.857

41. Pianezza, M. L., Sellers, E. M., & Tyndale, R. F. (1998). Nicotine metabolism defect reduces smoking. Nature, 393(6687), 750–750. 10.1038/31623

42. Plichta, M. M., Grimm, O., Morgen, K., Mier, D., Sauer, C., Haddad, L., Tost, H., Esslinger, C., Kirsch, P., & Schwarz, A. J. (2014). Amygdala habituation: a reliable fMRI phenotype. NeuroImage, 103, 383–390.

43. Poellinger, A., Thomas, R., Lio, P., Lee, A., Makris, N., Rosen, B. R., & Kwong, K. K. (2001). Activation and habituation in olfaction—an fMRI study. NeuroImage, 13(4), 547–560.

44. R.J. Reynolds Tobacco Company, et al. v. U.S. Food & Drug Administration, et al., 696 F.3d 1205, Nos. 11-5332, 12-5063 (U.S. Court of Appeals for the DC Circuit 2012).

45. R.J. Reynolds Tobacco Company, et al. v. U.S. Food & Drug Administration, et al., No. 6:20-cv-00176 (U.S. District Court for the Eastern District of Texas 2022).

46. R.J. Reynolds Tobacco Company, et al. v. U.S. Food & Drug Administration, et al., No. 24-189 (Supreme Court of the United States 2024a).

47. R.J. Reynolds Tobacco Company, et al. v. U.S. Food & Drug Administration, et al., 94 F.4th 863, No. 23-40076 (U.S. Court of Appeals for the Fifth Circuit 2024b).

48. R.J. Reynolds Tobacco Company, et al. v. U.S. Food & Drug Administration, et al., No. 6:20-cv-00176 (U.S. District Court for the Eastern District of Texas 2025).

49. Rankin, C. H., Abrams, T., Barry, R. J., Bhatnagar, S., Clayton, D. F., Colombo, J., Coppola, G., Geyer, M. A., Glanzman, D. L., Marsland, S., McSweeney, F. K., Wilson, D. A., Wu, C. F., & Thompson, R. F. (2009). Habituation revisited: an updated and revised description of the behavioral characteristics of habituation. Neurobiology of Learning and Memory, 92(2), 135–138.

50. Reiser, M., Yao, L., Wang, X., Wilcox, J., & Gray, S. (2017). A Comparison of bootstrap confidence intervals for multi-level longitudinal data using Monte-Carlo simulation. In D.-G. Chen & J. D. Chen (Eds.), Monte-Carlo simulation-based statistical modeling (pp. 367–403). Springer Nature.

51. Robinson, T. E., & Berridge, K. C. (1993). The neural basis of drug craving: an incentive-sensitization theory of addiction. Brain Research Reviews, 18(3), 247–291. 10.1016/0165-0173(93)90013-P

52. Rosenberg, H. (2009). Clinical and laboratory assessment of the subjective experience of drug craving. Clinical Psychology Review, 29(6), 519–534.

53. Sayette, M. A., Shiffman, S., Tiffany, S. T., Niaura, R. S., Martin, C. S., & Shadel, W. G. (2000). The measurement of drug craving. Addiction, 95(Suppl 2), S189–S210.

54. Shi, Z., Wang, A.-L., Fairchild, V. P., Aronowitz, C. A., Lynch, K. G., Loughead, J., & Langleben, D. D. (2023). Addicted to green: priming effect of menthol cigarette packaging on brain response to smoking cues. Tobacco Control, 32(e1), e45–e52. 10.1136/tobaccocontrol-2021-056639

55. Shi, Z., Wang, A.-L., Fairchild, V. P., Aronowitz, C. A., Padley, J. H., Lynch, K. G., Loughead, J., & Langleben, D. D. (2023). Effects of emotional arousal on the neural impact and behavioral efficacy of cigarette graphic warning labels. Addiction, 118(5), 914–924.

56. Shi, Z., Wang, A.-L., Liu, J., Audrain-McGovern, J., Lynch, K. G., Loughead, J., & Langleben, D. D. (2024). Delayed effects of cigarette graphic warning labels on smoking behavior. medRxiv, 2024.2001.2006.24300835. 10.1101/2024.01.06.24300835

57. Shiffman, S., Gwaltney, C. J., Balabanis, M. H., Liu, K. S., Paty, J. A., Kassel, J. D., Hickcox, M., & Gnys, M. (2002). Immediate antecedents of cigarette smoking: an analysis from ecological momentary assessment. Journal of Abnormal Psychology, 111(4), 531–545. 10.1037/0021-843X.111.4.531

58. Shiffman, S., West, R. J., & Gilbert, D. G. (2004). Recommendation for the assessment of tobacco craving and withdrawal in smoking cessation trials. Nicotine & Tobacco Research, 6(4), 599–614. 10.1080/14622200410001734067

59. So, J., Kim, S., & Cohen, H. (2017). Message fatigue: conceptual definition, operationalization, and correlates. Communication Monographs, 84(1), 5–29. 10.1080/03637751.2016.1250429

60. Tzourio-Mazoyer, N., Landeau, B., Papathanassiou, D., Crivello, F., Etard, O., Delcroix, N., Mazoyer, B., & Joliot, M. (2002). Automated anatomical labeling of activations in SPM using a macroscopic anatomical parcellation of the MNI MRI single-subject brain. NeuroImage, 15(1), 273–289.

61. U.S. Food and Drug Administration. (2011). Required warnings for cigarette packages and advertisements. Final rule. Federal Register, 76(120), 36628–36777.

62. U.S. Food and Drug Administration. (2022). Tobacco products; required warnings for cigarette packages and advertisements; delayed effective date. Federal Register, 87(40), 11295.

63. Vahratian, A., Briones, E. M., Jamal, A., & Marynak, K. L. (2025). Electronic cigarette use among adults in the United States, 2019–2023 (NCHS Data Brief, volume 524).

64. Wang, A.-L., Lowen, S. B., Romer, D., Giorno, M., & Langleben, D. D. (2015). Emotional reaction facilitates the brain and behavioural impact of graphic cigarette warning labels in smokers. Tobacco Control, 24(3), 225–232. 10.1136/tobaccocontrol-2014-051993

65. Wang, A.-L., Romer, D., Elman, I., Turetsky, B. I., Gur, R. C., & Langleben, D. D. (2015). Emotional graphic cigarette warning labels reduce the electrophysiological brain response to smoking cues. Addiction Biology, 20(2), 368–376. 10.1111/adb.12117

66. Wang, A.-L., Shi, Z., Fairchild, V. P., Aronowitz, C. A., & Langleben, D. D. (2019). Emotional salience of the image component facilitates recall of the text of cigarette warning labels. European Journal of Public Health, 29(1), 153–158.

67. World Health Organization. (2024). WHO global report on trends in prevalence of tobacco use 2000– 2030.

68. Wright, C. I., Fischer, H., Whalen, P. J., McInerney, S. C., Shin, L. M., & Rauch, S. L. (2001). Differential prefrontal cortex and amygdala habituation to repeatedly presented emotional stimuli. NeuroReport, 12(2), 379–383.

69. Zanto, T. P., Rubens, M. T., Thangavel, A., & Gazzaley, A. (2011). Causal role of the prefrontal cortex in top-down modulation of visual processing and working memory. Nature Neuroscience, 14(5), 656–661. 10.1038/nn.2773

